# The Influence of Hand Muscle Fatigue on Fine Motor Performance During Selected Activities of Daily Living in Healthy adults

**DOI:** 10.64898/2026.08.07.26359929

**Authors:** Mazin Alaa Alwash, Hossein Karimi

**Author notes:** Corresponding author. Mazin Alaa Alwash.

## Abstract

**Purpose:** By pairing repeated peripheral muscle fatigue induction with a functional and ADL-based assessment this study tests whether hand muscle fatigue alone is sufficient to impair fine motor performance in healthy adults performing ADL-inspired tasks.

**Participants and Methods:** Thirty healthy male and female university students performed 11 tasks twice—once in the pre-fatigue condition and once in the post-fatigue condition. The performance of each task was graded on a 0-4 scale. The score and the time to finish the task (TTFT) were recorded twice, for both the pre- and post-fatigue phases. Maximum force generation (MFG) of each participant grip was recorded prior to the tasks in the pre-fatigue phase and again in the fatigue condition after performing the fatigue protocol.

**Results:** Muscle fatigue did not have a significant effect on the fine motor performance of the thirty participants, neither (TTFT) nor the scores of each task (p > 0.05) (r=0.08). In contrast, hand muscle fatigue led to a significant decrease in the mean (MFG) for both males and females (p<0.01).

**Conclusion:** Hand muscle fatigue led to a significant decrease in the mean grip MFG for both sexes. however, this reduction did not translate into impaired fine motor performance during ADL-like tasks. Consequently, under muscle fatigue, hand grip changes during fine tasks tested are rare and have minor to no impact on hand performance. This study suggests that acute peripheral muscle fatigue can coexist with preserved fine motor performance in healthy populations.

## 1 INTRODUCTION

Muscle fatigue can be physically induced by muscular exertion and results in a reduction in the muscle’s ability to generate force. Muscles undergo fatigue due to repeated or prolonged activity. (Bestwick-Stevenson et al., 2022; Michael et al., 2012).

Fine motor skills of the hand are essential for activities of daily living (ADL) and many occupational tasks demand precision and sustained control. Acute peripheral muscle fatigue reaches initial recovery relatively quickly, often within approximately 2 minutes, during which the muscle’s maximum force generation (MFG) can return to its normal pre-fatigue state (Abaïdia et al., 2017). In contrast, chronic or central fatigue may persist and negatively affect cognitive resources and motor control.

Recent evidence has refined our understanding of how fatigue interacts with motor performance at the distal upper limb. A recent systematic review concluded that peripheral muscle fatigue does not consistently produce predictable impairments in hand and wrist performance, particularly during submaximal, precision-based tasks; the review highlighted the importance of fatigue specificity and motor abundance (redundant muscle contributions) as key moderators of performance outcomes (Forman et al., 2022b). In contrast, studies that induce mental (central) fatigue using prolonged cognitive tasks or mobile-app–based paradigms have reported selective impairments in fine motor accuracy, reaction time, and precision strength (Stafylidis et al., 2025). Despite this evidence, repeated muscle fatigue in an ADL-like context and occupation-relevant functional assessment adapted for healthy adults remain insufficiently investigated. By explicitly pairing repetitive peripheral-fatigue induction with functional and ADL-based assessment tasks with a scoring system that tests multiple hand grip patterns, this study tests whether hand muscle fatigue alone is sufficient to impair fine motor performance in healthy adults performing selected and ADL-inspired tasks.

## 2 PARTICIPANTS AND METHODS

Thirty participants (students of Istanbul Gelisim University’s) participated in the experiment: age: 22±1.45 years; height mean: 171±4.28cm weight: 68.69±7.66kg. All participants were right-side dominant. All participants had normal vision and no history of musculoskeletal injuries, neurological disorders, or cognitive dysfunctions. All participants acknowledged the experiment and gave consent prior to the experiment. The experiment took place at the occupational therapy laboratory at Istanbul Gelisim University.

### 2.1.1 Hand muscles fatigue measurement

Hand grip force was measured using the J00105 Lafayette Hand Dynamometer. Fatigue was defined as a 60% reduction from the recorded maximum grip force and this reduction reflects peripheral contractile failure. Standard verbal orders were used for motivation (“Push! … Push! … Relax”) were given to ensure maximal effort (Chengalur et al., 1990). Measurements were taken with the forearm in a functional position and the elbow at 90° flexion to prevent compensatory movements (Richards et al., 1996).

### 2.2 Tools and forms

Tools used for tests and tasks are similar to those used for Sollerman Hand Function Tests. Each tool is designed to assess one or more hand grips. See Table 1 Evaluation form and tools for reference.

**Table 1.**
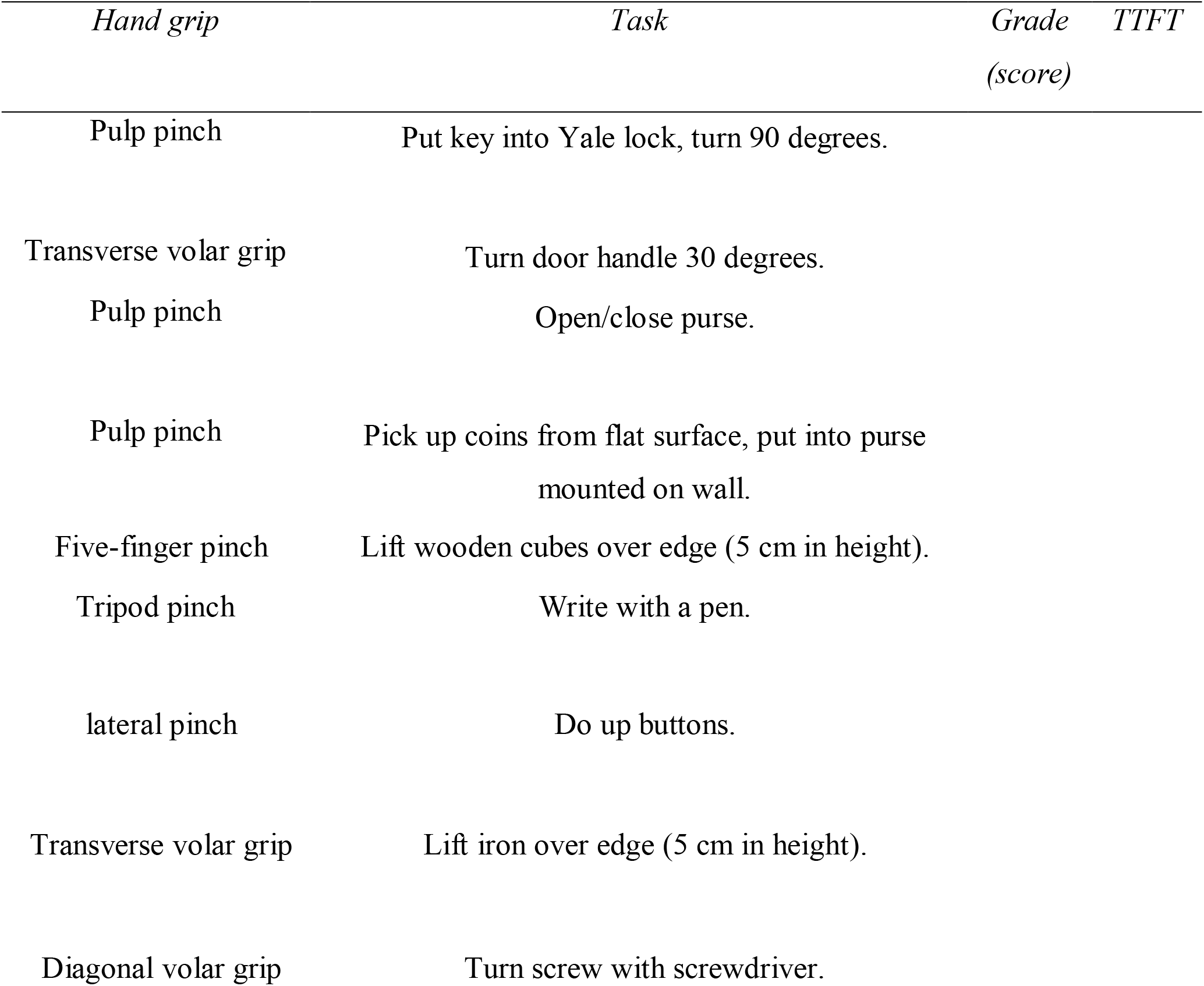
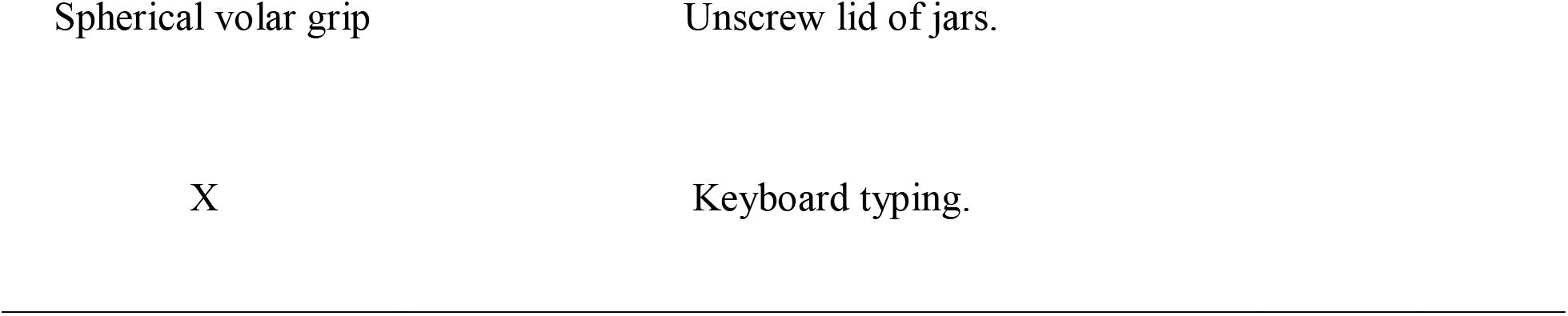
Evaluation form and tools.

The performance of each subject was graded from 0 to 4, with a maximum total score of 44 across the 11 tasks. The grading system considers task completion, execution time, level of difficulty, use of the prescribed grip pattern, and overall quality of hand performance. In the present study, prehension patterns was operationally defined as the participant’s ability to use the appropriate grip pattern for the performed task, and the subject’s performance was graded accordingly. See Table 2 The grading method for reference.

**Table 2.** The grading method.

2.3 Grading
| Score | Performance |
| --- | --- |
| 0 | The patient could not carry out the task |
| 1 | The task was partially performed within <u>60</u> seconds |
| 2 | The task was completed, but with great difficulty, or the task was not carried out with the prescribed hand-grip, or the task was not completed within <u>20</u> seconds but within <u>30</u> seconds |
| 3 | The task was completed, but with slight difficulty, or the task was carried out with the prescribed hand-grip but with slight divergence from normal, or the task was not completed within <u>10</u> seconds but within <u>20</u> seconds |
| 4 | The task was carried out without any difficulty within <u>10</u> seconds and with the prescribed hand-grip of normal quality. |

### 2.3 Grading

The Sollerman Hand Function Test was modified to better prioritize the assessment of fine motor skills. As the original test was designed for chronic conditions rather than acute, time-limited fatigue, tasks not primarily related to hand or fine motor function were excluded (from 20 tasks originally to 11 tasks), retaining only common ADL-based fine motor tasks while ensuring representation of different grip types. Additionally, the task time window was narrowed by 50% to reduce ceiling effects and increase sensitivity when assessing healthy individuals under fatigue, allowing performance to be captured before fatigue recovery occurred. See tables 1 and 2 above for reference.

#### Validity

We created a questionnaire that questions whether the test measures fine motor skills and grip functions and consulted 6 physical therapy and occupational therapy professors in Istanbul Gelisim University. The questionnaire has a scale from 1 to 10 for each task. Spearman rho (ρ) correlation was used, and the resulted correlation was strong (0.864 ρ) between the opinions of 6 professors.

Therefore, validity of the modified Sollerman was approved by the supervisor of the research and occupational and physical therapists of the College of Health Sciences of Istanbul Gelisim University.

### 2.4 Data Analysis Statistical analysis

All analyses were performed using IBM SPSS Statistics version 26. Normality of continuous variables was assessed visually (histograms and Q–Q plots) and by the Shapiro–Wilk test.

Because several outcome variables showed non-normal distributions and large inter-individual variability, nonparametric statistics were used. The significance level was set at p < 0.05.

#### Normalization of TTFT

To make task responses comparable across tasks with different baseline durations, TTFT values were normalized within-subject as percent change from baseline using the following formula:

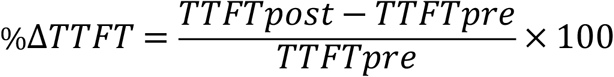

Normalized TTFT is the percentage change of time to finish the task, this ensures every task is equal in compression and prevents heterogeneity. A participant-level %ΔTTFT mean was calculated as the arithmetic mean of all available tasks %ΔTTFT values for each participant. This single summary value per participant was used for the global (primary) inferential test to avoid pseudoreplication.

#### Reliability (internal consistency)

Internal consistency of the Modified-Sollerman hand function test was assessed using Cronbach’s alpha (α) computed on the TTFT item set (22 items). Cronbach’s α resulted in (α =0.625) commonly considered moderate.

#### Primary analysis (fatigue effect)

The primary inferential analysis evaluated whether the participant-level mean percentage change in time to finish the task (%ΔTTFT) differed from zero. Because the distribution of %ΔTTFT deviated from normality, a two-tailed one-sample Wilcoxon signed-rank test was used to test the null hypothesis that the median participant-level mean %ΔTTFT was equal to zero. Statistical significance was set at p < 0.05. Performance scores for each of the 11 tasks (rated on a 4-point scale) were compared between pre- and post-fatigue conditions using the Wilcoxon signed-rank test. Task-level analyses were exploratory and intended to characterize the direction and magnitude of fatigue effects across individual tasks. Consequently, interpretation of these secondary analyses emphasized effect sizes and confidence intervals rather than strict dichotomous statistical significance, and no corrections for multiplicity were applied.

#### Bayesian analysis

As a complementary analysis, a Bayesian one-sample normal model was performed on the participant-level mean %ΔTTFT using diffuse priors for the mean and variance. Bayes factors (BF_01_) together with 95% posterior credible intervals were reported to quantify evidence supporting the null hypothesis relative to the alternative hypothesis.

#### Effect size

Effect sizes for Wilcoxon signed-rank tests were quantified using the standardized effect size (r), calculated as the standardized test statistic divided by the square root of the number of observations 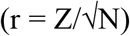, where appropriate. Effect sizes were interpreted according to Cohen’s guidelines (0.10 = small, 0.30 = medium, 0.50 = large).

### 2.5 Procedure

The experiment consisted of three phases: pre-fatigue, fatigue induction, and post-fatigue

#### 2.5.1 Pre-fatigue phase

Starts by measuring the highest MFG, then the subjects started to perform 11 tasks of the Modified Sollerman test in a normal (pre-fatigue) state. After each task completion the subject’s performance was recorded. When the participant finished the total 11 tasks, the fatigue induction protocol began.

Figure 1 above illustrate the procedure process

**Figure 1.**
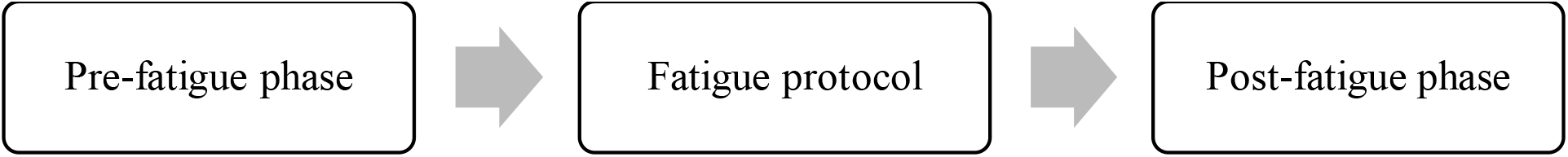
the procedure

#### 2.5.2 Fatigue induction protocol

The protocol consisted of three fatigue-induction bouts separated by two rest periods. Muscle fatigue was induced using a hand–finger flexion resistive spring device. Fatigue was defined as a 60% reduction in the total MFG, after which a rest period was provided. We ensured that the MFG returned to 90% of total value before inducing fatigue again. This process was repeated until the 3rd fatigue induction was completed, after which the next post-fatigue phase began.

Figure 2 below illustrate the fatigue protocol

**Figure 2.**
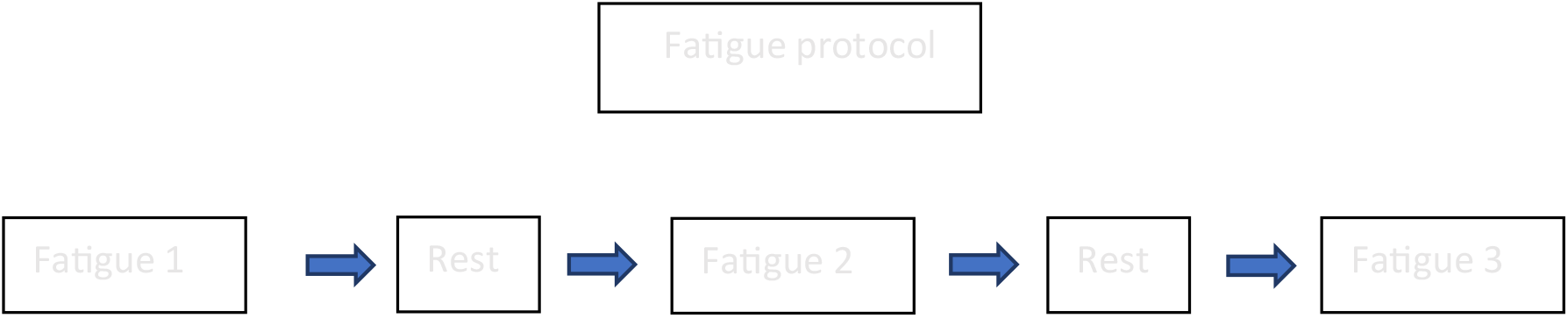
the fatigue induction protocol

#### 2.5.3 Post-fatigue phase

After completing the fatigue protocol, the subject immediately began the post-fatigue phase, performing the same 11 tasks in a fatigued state. The TTFT and performance scores were recorded on a separate sheet to prevent the examiner from seeing previous results. After completing the first six tasks, fatigue was induced for the fourth and final time using the same procedure. The experiment concluded after the subject performed the eleventh and final task in the post-fatigue phase.

Figure 3 below illustrate the post fatigue phase process.

**Figure 3.**
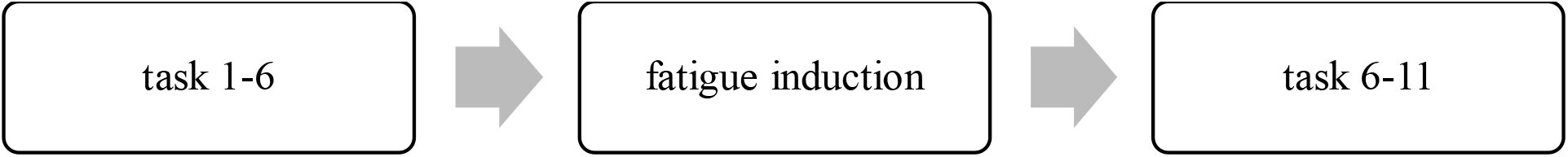
post fatigue phase

## 3 RESULTS

The (MFG) mean was 26.31±6.53 kg force for male samples (23 participants) and 13.75±3.88 kg force for females (7 participants).

The highest MFG recorded was 22 kg for females and 39 kg for males. In the post-fatigue phase, the MFG showed a highly significant decrease after performing the fatigue protocol for females, decreasing by 68.21%; the mean was 4.37±2.5 kg force, and a 65.64% decrease in the MFG for males, with a mean of 9.04±2.57 kg force.

Wilcoxon signed-rank test comparing participants’ mean normalized TTFT to 0 revealed no significant fatigue-related change (p = 0.629). The effect size was very small (r=0.08).

Furthermore, the Hodges–Lehmann estimate of the median difference in normalized TTFT was 0.93%, with a 95% confidence interval of [–3.23%, 5.28%]. This interval includes zero, indicating no significant median shift.

Bayesian one-sample normal model was performed on the participant-level mean normalized TTFT (%ΔTTFT) using diffuse priors. The analysis yielded a Bayes factor of BF_01_ = 6.69, indicating that the observed data were approximately 6.7 times more likely under the null hypothesis than under the alternative hypothesis, providing moderate evidence for the absence of a global fatigue effect. The posterior 95% credible interval ranged from −4.62% to 3.34%, further supporting that the overall fatigue-related change in normalized TTFT was centered near zero.

Task-level analysis showed that task 1 (pulp pinch grip) exhibited the largest effect (p = 0.011) with medium effect size (r = 0.459). In contrast, the remaining 10 tasks demonstrated small or trivial effects (p > 0.05).

The Wilcoxon signed-rank test was used on the samples’ scores and showed no significant difference (p > 0.05) between the pre- and post-fatigue values.

## Discussion

Since many occupations require fine control of small objects, repeated finely controlled tasks may lead to hand muscles fatigue. This study aimed to assess how the hand would function under fatigue while undergoing certain daily living inspired tasks. The authors acknowledge that the Sollerman Hand Function Test was not originally designed to be used with healthy individuals. Furthermore, this test cannot easily assess fatigue, primarily due to the time restrictions imposed by fatigue recovery. Muscles recover relatively quickly, typically within about 2 minutes. Therefore, the subject would not be able to complete all 11 tasks without sufficient recovery. To address this limitation….

1. A task-selected version of the Sollerman Hand Function Test was employed by narrowing the task time window and selecting ten tasks that are the most relevant to common activities of daily living. This approach was adopted to reduce ceiling effects and account for the rapid recovery and high functional performance typically observed in healthy individuals. The task-selected version demonstrated moderate internal consistency.
2. A muscle fatigue protocol was implemented to induce muscle fatigue at four predefined intervals throughout the experiment. Fatigue status was objectively verified by measuring grip force using a handheld dynamometer, ensuring that participants remained in a fatigued state while performing the tasks.

The experiment resulted in MFG dropping significantly (p<0.01) in the post-fatigue phase compared to the pre-fatigue phase in both genders. while the MFG has been measured multiple times after each fatigue during the fatigue protocol, in the third fatigue, the MFG was usually the lowest and took the most time to recover and this supports previous studies that tested fatigue effect on hand grip MFG (Mahdavi et al., 2021) (Kumar, 2006).

Comparing participants’ mean normalized TTFT to zero revealed no significant fatigue-related change (p = 0.629), with a very small effect size (r = 0.08), suggesting that performing the selected ADL-inspired submaximal tasks under muscle fatigue had near zero effects on task completion time, with rare grip alterations observed during performance suggesting almost normal prehension patterns even after fatiguing protocol. These findings are consistent with a recent systematic review reporting that muscle fatigue does not produce consistent impairments in performance outcomes, particularly during submaximal functional tasks (Forman et al., 2022b). The findings also well align with the notion that the magnitude of muscle fatigue does not directly determine performance outcomes and does not necessarily result in task failure during familiar activities of daily living, where compensatory strategies may help preserve task performance (Forman et al., 2022b; Enoka & Duchateau, 2008). One possible explanation is neuromuscular adaptability, whereby the central nervous system adjusts motor output in response to fatigue. This adaptability may be facilitated by motor redundancy, which allows the redistribution of load among synergistic muscles capable of contributing to the same task. As discussed by Enoka and Duchateau (2008), motor performance depends on the flexibility of the neuromuscular system to reorganize coordination patterns rather than solely on the force-generating capacity of individual muscles.

Prehension patterns of the subjects was very high and the subjects mostly used the correct grip for the specific task despite being under muscle fatigue. However, most of the grip alterations were observed in two tasks, particularly the screwdriver (diagonal volar grip) and buttoning (lateral pinch) tasks, but these changes were associated with only brief temporal delays and did not significantly affect TTFT or lead to task failure. This may be explained by the principles of motor redundancy and prehension synergies. Previous research has demonstrated that the central nervous system can achieve the same task goal through multiple interdigit coordination patterns, reflecting the inherent redundancy of the hand system (Zatsiorsky & Latash, 2004). Although these studies primarily investigated finger-level coordination and force redistribution rather than transitions between clinically defined grip patterns, they demonstrate that fatigue-related adaptations in coordination can occur without necessarily compromising task success (Singh et al., 2014). Alternatively, evidence from hand muscle EMG studies suggests that performance may also be preserved through increased neural drive and intermuscular synchronization while maintaining stable muscle coordination patterns throughout fatigue (Danna-Dos Santos et al., 2010). Together, these findings further support that fatigue does not necessarily impair functional hand performance despite alterations in motor control.

The present study included a relatively limited sample size (n = 30). Although no statistically significant differences were observed in fine motor performance following fatigue, the observed effect size was very small (r = 0.08). Nevertheless, the modest sample size may have limited statistical power to detect small effects. Future studies with larger samples and a more balanced sex distribution are recommended to further investigate the relationship between hand muscle fatigue and fine motor performance. The modified scoring system inter-rater reliability and comprehensive psychometric validation should be established before broader application.

## 4 Conclusion

This study investigated the effect of hand muscle fatigue on performance during selected ADL-inspired fine motor tasks in healthy individuals. The fatigue protocol resulted in a significant reduction in maximum grip force in both males and females; however, this reduction was not accompanied by a significant increase in task completion time or deterioration in prehension patterns, as assessed using the modified Sollerman scoring system. These findings suggest that, under the fatigue protocol and outcome measures used in this study, hand muscle fatigue did not produce significant impairments in task completion time or prehension patterns during the selected submaximal ADL-inspired tasks.

## Data availability

the research data supporting the findings of this study are available from the corresponding author upon reasonable request.

The ethical committee of Istanbul Gelisim University approved the experimental procedures.

## approval number

2023-04-142

date: 19.4.2023

